# Bacterial Etiology of Urinary Tract Infections In patients treated at Kenyan Health Facilities And their resistance towards commonly used antibiotics

**DOI:** 10.1101/2022.10.25.22281521

**Authors:** Susan Kiiru, John Maina, Japhet Katana, John Mwaniki, Benon B Asiimwe, Stephen E Mshana, Katherine Keenan, Stephen H. Gillespie, John Stelling, John Kiiru, Matthew T G Holden, HATUA Consortium

## Abstract

1.0

**Background:** Evidence-based empirical antibiotic prescribing requires knowledge of local antimicrobial resistance patterns. The spectrum of pathogens and their susceptibility strongly influences guidelines for empirical therapies for urinary tract infections (UTI) management.

**Objective:** This study aimed to determine the prevalence of UTI causative bacteria and their corresponding antibiotic resistance profiles in in three counties of Kenya. Such data could be used to determine the optimal empirical therapy.

**Methods:** In this cross-sectional study, urine samples were collected from patients who presented with symptoms suggestive of UTI in the following healthcare facilities; Kenyatta National Hospital, Kiambu Hospital, Mbagathi, Makueni, Nanyuki, Centre for Microbiology Research, and Mukuru Health Centres. Urine cultures were done on Cystine Lactose Electrolyte Deficient (CLED) to isolate UTI bacterial etiologies, while antibiotic sensitivity testing was done using the Kirby-Bauer disk diffusion using CLSI guidelines and interpretive criteria.

**Results:** A total of 1,027(54%) uropathogens were isolated from the urine samples of 1898 participants. Staphylococcus spp. and Escherichia coli were the main uropathogens at 37.6% and 30.9 %, respectively. The percentage resistance to commonly used drugs for the treatment of UTI were as follows: trimethoprim (64%), sulfamethoxazole (57%), nalidixic acid(57%), ciprofloxacin (27%), amoxicillin-clavulanic acid (5%), and nitrofurantoin (9%) and cefixime (9%). Resistance rates to broad-spectrum antimicrobials, such as ceftazidime, gentamicin, and ceftriaxone, were 15%, 14%, and 11%, respectively. Additionally, the proportion of Multidrug-resistant (MDR) bacteria was 66%.

**Conclusion:** High resistance rates toward fluoroquinolones, sulfamethoxazole, and trimethoprim were reported. These antibiotics are commonly used drugs as they are inexpensive and readily available. Based on these findings, more robust standardised surveillance is needed to confirm the patterns observed while recognizing the potential impact of sampling biases on observed resistance rates.

## Background

Urinary tract infections (UTIs) are among the community’s most common infections [1] accounting for nearly 25% of all common infections[2]. Globally, UTI is more common among the aged, prepartum neonates, pregnant women, and hospitalized patients, especially those in Intensive Care Units (ICU) and those with indwelling catheters[3]. Usually, community-acquired UTIs are more prevalent than hospital-acquired UTIs[4]. Even though UTIs can be treated using antibiotics, previous widespread antibiotic usage without proper susceptibility testing has inevitably led to an increase in the proportion of UTI pathogens resistant to affordable and available antibiotics. Understanding local antimicrobial resistance patterns is a baseline step for evidence-based empirical antibiotic prescribing[5]. Continuous surveillance and monitoring of AMR trends are vital to informing clinical decisions during empirical management of UTIs in health facilities lacking laboratory capacity to perform culture and sensitivity tests. In order to administer an appropriate empirical therapy, it is critical to know the main uropathogens. [6].

It is estimated that ∼50% of women will have at least one UTI episode in their lifetime, while 20-40% will have recurrent episodes [7]. Pregnant women are more susceptible to UTIs due to hormonal and physiologic changes predisposing them to bacteriuria [8]. Up to 70% of pregnant women develop glycosuria, which encourages bacterial growth in the urine [9]. Antibiotic therapy for 70% of these infections usually begins before microbiological test results are known[10]. Furthermore, empirical therapy without a pretherapy urine culture is often used in women with acute uncomplicated cystitis. The rationale for this approach is based on the highly predictable spectrum of etiologic agents causing UTIs and their antimicrobial resistance patterns. However, antimicrobial resistance among uropathogens causing complicated and uncomplicated community-acquired UTIs is gradually increasing. Most significant has been the increasing resistance to trimethoprim/sulfamethoxazole (co-trimoxazole), commonly sold as Septrin^(tm)^, the current drug of choice for treating acute uncomplicated cystitis in women [11]. In addition, SXT is frequently associated with concurrent resistance to other antibiotics classes, resulting in multidrug-resistant uropathogens [11].

In Kenya, the most common Gram-negative etiologic agents reported in UTI cases in order of decreasing prevalence are *Escherichia coli, Klebsiella, Enterobacter*, and *Proteus*. At the same time, *Staphylococcus* is the most common Gram-positive genus [12] followed by *Enterococcus*. Determining microorganisms and their antibiotic sensitivity patterns allows for good treatment outcomes, controls the increase of antimicrobial prescription, and helps prevent antimicrobial resistance, a public health problem worldwide. This paper establishes the common uropathogens from different healthcare facilities across Kenya and their corresponding susceptibility patterns.

## Methodology

### Study Overview

The finding reported in the present manuscript is a subset of a large Holistic Approach To Unravel Antibiotic-resistance (HATUA) consortium study in East Africa https://pubmed.ncbi.nlm.nih.gov/34006022/. This East African consortium included Kenya, Uganda, and Tanzania. The study employed social science, microbiological, and molecular biology disciplines to unravel drivers of AMR holistically using UTI as the flag disease. In Kenya’s HATUA chapter, the study was conducted in Nairobi, Makueni, and Nanyuki metropolis regions.

### Study design and site

A cross-sectional study design was employed to establish the common uropathogens recovered from purposively recruited 1898 patients with UTI-like symptoms. The participants were recruited from major public hospitals in Nairobi, Nanyuki, and Makueni counties in the former Nairobi, Central, and Eastern provinces, respectively, as shown in Fig 1. These counties were selected as there was limited information on uropathogens and their antibiotic resistance profiles. In each region, sampling was done from one level 5 hospital and in a maximum of 3 smaller hospitals or clinics (health centers, or Level 3 or 4), among participants who were living within the 70 km radius of the recruitment hospitals/clinics. The sampling regions included; Kenyatta National Hospital (a public, teaching, and national referral level 6 hospital), Mbagathi County level 5 Hospital (referral hospital in Nairobi region), Kiambu County level 5 Hospital (a referral hospital in Nairobi region), Nanyuki Teaching and Referral level 5 Hospital (referral hospital in the Central Region), and Makueni County Referral Level 5 Hospital (referral hospital in the Eastern).

**Fig 1:**
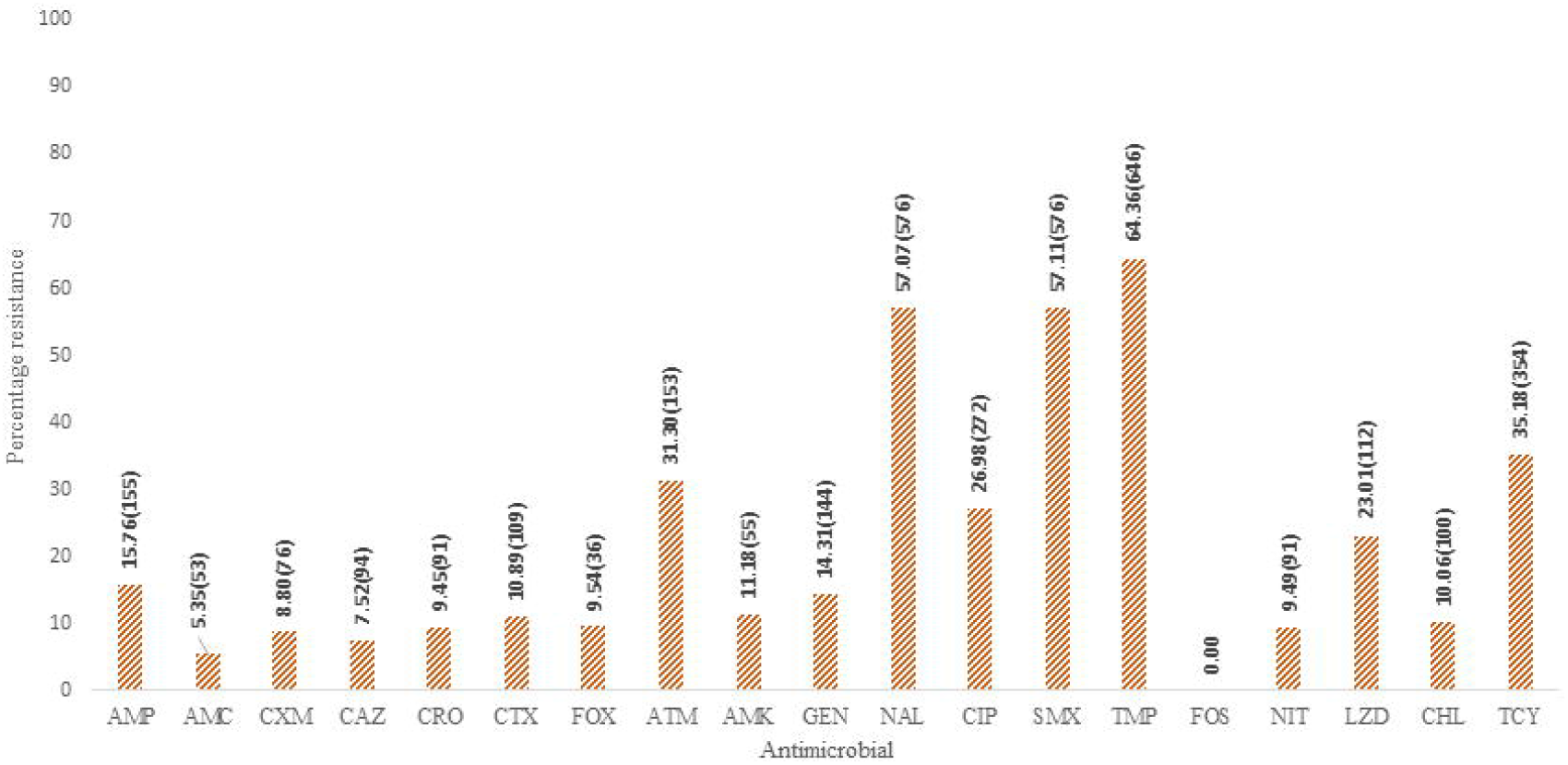
The location of study areas from where the recruitment occurred.

Most patients are referred from other facilities or come as self-referrals after treatment has been initiated at lower-level facilities. Mary Mother Mission Mukuru Healthcare (a health clinic in the second largest informal settlement in Nairobi), Ruiru Family Clinic (a private clinic in Nairobi county), KEMRI Centre for Microbiology Research Lab (a referral research facility that also houses the laboratory offering food handlers medical certification in Nairobi region).

### Ethical Statement

Before the start of the study, ethical approval was obtained from the Scientific Ethical Review Committee (SERU) of Kenya Medical Research Institute (No. KEMRI/SERU/CMR/P00112/3865). Additional ethical approval was obtained from National Commission for Science, Technology, and Innovation (NACOSTI) and the study recruitment healthcare facilities. Consent was sought before recruiting the participants for the study, and the names of the participants were not taken. The results approved by the project Principal Investigator were sent back to the medical doctor weekly, and presentations were done during the hospital’s Continuing Medical Education meetings.

### Recruitment and laboratory procedures

#### Recruitment and sampling

A medical doctor (resident in the hospital selected for study) identified patients who had symptoms suggestive of UTIs like burning pain after urinating, urgent need to urinate, pain or pressure in the lower abdomen, and cloudy, dark, bloody, or strange-smelling urine. The patients meeting the inclusion criteria were then referred to a study recruiting healthcare worker who was in a separate room within the hospital. For the inpatients, the doctor would request urine culture, and the nurses would assist in collecting the clean catch mid-stream urine aseptically, then send the urine to the hospital laboratory. The researcher in the laboratory would then request the nurse to help identify these inpatients and invite them to be recruited in the study, where they signed or thumb-signed the consent form. For children inpatients, the parent or guardian would give consent for them to participate in the study, then the child would thumb sign the assent form.

The researcher introduced the patient to the study and briefly explained the study objectives. Willing patients were then taken through the study informed consent document (ICD) in a language they could understand (English and/or Kiswahili), where they signed the ICD, and for those who couldn’t write, they thumb printed the ICD. Those who consented were given a study number that was filled in their questionnaire form. This study number was also written on the sample collection container. After consenting, the participant was requested to submit the urine samples.

A purposive sampling method was used to recruit patients that presented with UTI-like symptoms in the eight recruitment sites. Considering that there is overwhelming evidence that UTI diagnosis is complex and based mainly on dipsticks and microscopy that are frequently inaccurate[13], and considering that proper cultures for UTI are rarely performed in order to confirm UTI cases in many hospitals[8], therefore, the recruitment was standardised across countries, as per the protocol[14] briefly, to estimate precision, under a binomial model, the numbers required to obtain a 95% CI for the prevalence of 0.5 with width no greater than 0.1 would be a little under 400 (384). That model relies on there being no underlying population or sampling structure and so will lead to an underestimate of the true required numbers in our complex study. Our larger study size of 600 per country provided some robustness to our ability to estimate this parameter with the desired accuracy while allowing us to uncover some of the population structures that, if modelled correctly, will improve the precision in our estimate of prevalence. In level 2, 3, 4 and 5 hospitals in each study area, we recruited adult and child outpatients (minimum of 90% of the total sample) that a doctor identified as suffering with UTI-like symptoms (eg, burning/irritation during urination, dysuria and pyuria). In level 5 hospitals, we also recruited inpatients (maximum of 10% of the total). For non-pregnant child patients aged under 18 years, data was provided by an accompanying parent or guardian. Our sample is representative only of the population of clinic attendees rather than the general population and is likely to include a higher proportion of patients with treatment failures who are wealthier and patients living closer to clinics. However, clinic attendees are an important patient subset as these are the individuals specifically for whom clinicians must make patient management and treatment decisions. The distribution of participants across the 8 recruitment sites depended on the proportion of patients visiting the hospital presenting with UTI-like symptoms and hence were not proportionally distributed.

#### Urine collection procedure

Clean-catch mid-stream urine was collected into 20 mL calibrated sterile screw-capped universal bottles. The adult participants were guided on how to collect the specimen by the fieldworker(s) aseptically. The fieldworker also assisted parents in collecting urine specimens from young children. Briefly, the parents had to take care the children did not touch the perineum with the collection tube; children older than two years who were able to follow instructions from their parents provided a mid-stream or clean catch sample of urine directly into sterile urine bottles under observation by one of the research staff. For those younger than two years or those who could not follow instructions, an in-and-out catheter was put in, and a sterile feeding tube size 5 or 6 was used to collect urine. A unique study identification number and barcode label was applied to the sample immediately after urine collection. To ensure urine was not contaminated by dipstick before culture, the collected urine was aliquoted in another sterile urine container labeled with the patient’s unique number. The dipstick was then put in one of the aliquots for testing. The other portion of the urine aliquot was then kept in a cool box to be transported to the microbiology laboratory for further processing within 4 hours.

#### Bacterial isolation and identification

Urine samples were analyzed using dipstick strips, and urine colony count was determined in colony-forming units (CFUs) by culture as the gold standard. The patients were first screened by dipstick analysis in the hospital, using nitrites and leukocyte esterase parameters to rule out the positive and negative tests. Both positive and negative urines by dipstick were then cultured on Cystine Lactose Electrolyte Deficient (CLED) agar and incubated at 37 □ overnight. Culturing both negative and positive urine was to detect possible UTI cases that may have been missed by dipstick test screening. A specimen was considered positive for Urinary Tract Infection (UTI) if the organism colony count was determined to be ≥10^4^CFU/mL with >5 pus cells per high-power field observed on microscopic examination of the urine as described by [15]. Additionally, bacterial identification was made using colony morphology, Gram staining, Triple Sugar Iron (TSI), citrate test, Lysine Indole Motility (LIM), Methyl Red and Voges Proskauer (MRVP), urease test, oxidase, catalase, and coagulase tests using commercially available NCTC 13420 to help in identification of *Acinetobacter baumannii*, NCTC 10975 to identify *Proteus mirabilis*, NCTC 12028 to identify *Morganella moganii*, NCTC 8900 to identify *Serratia marcescens, E*.*coli* ATCC 25922 and *S. aureus* ATCC 25923.

#### Antimicrobial Susceptibility testing

Antimicrobial drug susceptibility testing was performed by the Kirby Bauer disk diffusion method on Mueller–Hinton agar following the M02 CLSI 2019 guidelines. The following Oxoid(tm) antimicrobial agents were used; ampicillin (AMP 10µg), amoxicillin/clavulanic acid (AMC 20/10µg), cefoxitin (FOX 30µg), cefotaxime (CTX 30µg), ceftazidime (CAZ 30µg), cefixime (CXM 30µg), ceftriaxone (CRO 30µg), ciprofloxacin (CIP 5µg), nalidixic acid (NAL 30µg), nitrofurantoin (NIT 300µg), trimethoprim(TMP 5µg), sulfamethoxazole (SMX 200-300 µg), gentamicin (GEN 10µg), chloramphenicol (CHL 30µg), tetracycline (TCY 30µg), linezolid(LNZ 30µg), erythromycin(ERY 30µg), fosfomycin(FOS 30µg), amikacin (AMK 30µg), aztreonam (ATM 30µg). For each Gram-negative isolate, two plates with antibiotics were used, labeled as plates A and B. Plate A was used to screen for potential Extended Spectrum Beta-Lactamase (ESBL) production. The arrangement of antibiotics was as follows, penicillins (AMP), 3^rd^ generation cephalosporins (CXM, CRO, CAZ, CTX), cephamycin (FOX), monobactam (ATM), beta-lactamase inhibitor at the middle (AMC), and a 4^th^-generation cephalosporin (FEP). In plate B, CIP, NAL (targeting quinolones and fluoroquinolone resistance), GEN, AMK, and STR (targeting aminoglycoside resistance), CHL, TMP, SMX, FOS, NIT, and TCY were used since they are used in hospitals for treatments. Gram-positive isolates had two plates labeled plates A and B. Plate A had AMP, AMC, FOX, TMP, GEN, and NIT, and B had SMX, CIP, NAL, CHL, TCY, linezolid, erythromycin, and fosfomycin. *Escherichia coli* ATCC 25922 and *Staphylococcus aureus* ATCC 25923 were used for quality control of media quality and disc potency. The antibiograms generated were then used to cluster the isolates into various resistance profiles ranging from fully sensitive to multidrug-resistant. The strains exhibiting resistance to 3 or more classes or subclasses of antibiotics were scored as MDR[16].

## Results

### Socio-demographic Characteristics and Prevalence of UTI

A total of 1898 patients who presented with UTI-like symptoms were recruited between May 2019 and August 2020. Among them, 1546 (81.5%) were females, while 352 (18.5%) were males. A total of 67 (3.5%) were children (≤17 years), and 1831 (96.5%) were adults. Of the adults, 1708 (93.3%) were outpatients, while 123 (6.7%) were inpatients. Twenty-two (32.8%) of the children were inpatients, while 45 (67.2%) were outpatients, as illustrated by Table 1 below. The average age of the study participants was 30.7 ± 12.1 years, with the youngest being one year old and the oldest 102 years.

**Table. 1.** Demographic characteristics of the study participants.

| Social demographic characteristics |  | Female<br>N (%)<br>(n = 1546) | Male<br>N (%)<br>(n = 352) | Total<br>(n = 1898) |
| --- | --- | --- | --- | --- |
| Patient type | Adult inpatient | 81 (65.3) | 43 (34.7) | 124 (100) |
|  | Adult outpatient | 1429 (83.7) | 278 (16.3) | 1707 (100) |
|  | Child inpatient | 9 (40.9) | 13 (61.9) | 22 (100) |
|  | Child outpatient | 28 (62.2) | 17 (37.8) | 45 (100) |
| Recruitment site | CMR | 17 (58.6) | 12 (41.4) | 29 (100) |
|  | Kenya Ruiru Family clinic(KRF) | 22 (81.5) | 5 (18.5) | 27 (100) |
|  | KIAMBU | 182 (78.4) | 50 (21.6) | 232 (100) |
|  | KNH | 84 (67.2) | 41 (32.8) | 125 (100) |
|  | MAKUENI | 354 (92.7) | 28 (7.3) | 382 (100) |
|  | MBAGATHI | 112 (73.7) | 40 (26.3) | 152 (100) |
|  | MMM | 320 (82.7) | 67 (17.3) | 387 (100) |
|  | NANYUKI | 413 (85.5) | 70 (14.5) | 483 (100) |
| Study Location | Central Kenya (Nanyuki) | 414 (85.4) | 71 (14.6) | 485 (100) |
|  | Larger Nairobi Metropolis | 778 (75.5) | 253 (24.5) | 1031 (100) |
|  | MAKUENI | 354 (92.9) | 27 (7.1) | 381 (100) |
| Age group | < 5 years | 10 (33.3) | 20 (66.7) | 30 (100) |
|  | 6-18 years | 66 (81.5) | 15 (18.5) | 81 (100) |
|  | 19-24 years | 425 (92.6) | 34 (7.4) | 459 (100) |
|  | 25-30 years | 497 (84.5) | 91 (15.5) | 588 (100) |
|  | 31-35 years | 249 (83.3) | 50 (16.7) | 299 (100) |
|  | 35-40 years | 142 (78.5) | 39 (21.5) | 181 (100) |
|  | 41-45 years | 60 (69.0) | 27 (31.0) | 87 (100) |
|  | 46-50 years | 27 (60.0) | 18 (40.0) | 45 (100) |
|  | 51-55 years | 21 (65.6) | 11 (34.4) | 32 (100) |
|  | 56-60 years | 14 (53.8) | 12 (46.2) | 26 (100) |
|  | > 60 years | 35 (50.7) | 34 (49.3) | 69 (100) |

The overall pathogen isolation from patients with UTI-like symptoms was 1027(54.1 %), with women having a higher isolation rate of 917 (59.3%) compared to men at 110(31.3%). Bacteriuria was highest in the age group 25-30 years at 16.2% and the lowest (1.2%) in children under five years.

### Bacterial isolates

Of all (1027/1898) UTI-positive patients, *Staphylococcus* spp. and *E. coli* were the main uropathogens isolated at 37.6% and 30.9%, respectively. Other uropathogens included *Enterococcus* spp 86 (8.4%), *Klebsiella* spp 91 (8.9%), *Proteus* spp 69(6.7%), *Pseudomonas* spp. 8 (0.8%), *Acinetobacter* spp. 8 (0.8%). A total of 62(6%) organisms were regarded as miscellaneous in this study, as shown in Fig1. Gram-positive uropathogens were more prevalent at 524 (51%), followed by Gram-negative organisms at 490 (48%) and yeast at 10 (1%.)

### Antimicrobial Susceptibility Testing

The recovered uropathogens were subjected to antibiotic susceptibility testing.

However, yeast isolates were not tested, resulting in 1016 bacterial isolates for AST. Overall, the percentage resistance to commonly used UTI drugs were as follows; trimethoprim (64%), sulfamethoxazole (57%), nalidixic acid (57%), ciprofloxacin (27%), nitrofurantoin (9%), cefixime (9%) and amoxicillin-clavulanic acid (5%). Resistance rates to broad-spectrum antimicrobials, such as gentamicin, ceftriaxone, and ceftazidime, were at 14%, 9%, and 8%, respectively. Cefixime, nitrofurantoin, and amoxicillin/clavulanic acid were the most effective agents against these isolates, while trimethoprim, sulfamethoxazole, and nalidixic acid were the most antibiotics with the highest resistance rates as seen in Fig 3. *Staphylococcus* spp., the dominant uropathogen, showed the highest resistance towards trimethoprim (76%), nalidixic acid (70%), sulfamethoxazole (67%), and ampicillin (64%). Similarly, resistance towards cefoxitin was 38%, signifying the possibility of methicillin-resistant *S. aureus* (MRSA) was 38%. Moreover, all isolated *Pseudomonas* spps (8) showed resistance towards cefuroxime (100%) as well as high resistance towards ciprofloxacin (39%), nalidixic acid (50%), nitrofurantoin (38%), sulfamethoxazole (38%) and trimethoprim (38%). The proportion of multidrug-resistant (MDR) pathogens was 66%, with the most common MDR phenotype observed being a combination of nalidixic acid, sulfamethoxazole, and trimethoprim. Nalidixic acid is not included in the ECDC definition of MDR, but because it is routinely used for UTI treatment, modifications were made to the definition of MDR. The resistance towards nalidixic acid was high across all the recruitment sites, ranging from 52 to 71%, as Table 3 depicts. The resistance towards the mainstay treatments of UTI - beta-lactams, quinolones, fluoroquinolones, and aminoglycosides, commonly abbreviated as BFQA - was high in Kenyatta National Hospital, ranging from 31 to 43% and was lowest at Ruiru Family Clinic ranging from 0 to 13%, as shown in the highlighted part of Table 3.

**Table 3:** Antibiotic Resistance % across all recruitment sites.

| Study Site | AMP | AMC | CXM | CAZ | CRO | CTX | FOX | ATM | AMK | GEN | NAL | CIP | SMX | TMP | FOS | NIT | LNZ | CHL | TCY |
| --- | --- | --- | --- | --- | --- | --- | --- | --- | --- | --- | --- | --- | --- | --- | --- | --- | --- | --- | --- |
| CMR | 41(17) | 18(17) | 0 | 18(17) | 18(17) | 24(17) | 18(17) | 33(6) | 0 | 24(17) | 71(17) | 29(17) | 41(17) | 47(17) | 0 | 6(20) | 0 | 12(17) | 29(35) |
| Kenyatta National Hospital | 39(75) | 32(75) | 41(29) | 37(75) | 36(72) | 37(75) | 43(75) | 63(35) | 43(35) | 37(75) | 71(75) | 37(75) | 61(75) | 64(75) | 3(35) | 19(77) | 34(35) | 20(75) | 47(21) |
| Kiambu County hospital | 23(126) | 21(126) | 37(41) | 20(126) | 21(125) | 21(126) | 25(126) | 50(62) | 10(62) | 17(126) | 61(126) | 22(126) | 51(127) | 56(127) | 0 | 12(125) | 33(34) | 10(130) | 30(33) |
| Makueni Level 5 | 39(265) | 13(265) | 15(265) | 15(265) | 17(265) | 18(265) | 17(265) | 47(135) | 10(265) | 11(265) | 55(265) | 20(265) | 51(250) | 69(250) | 0 | 6(20) | 19( 6) | 10(130) | 43(27) |
| Mbagathi hospital | 26(34) | 26(34) | 33(12) | 18(34) | 21(33) | 26(34) | 24(34) | 53(15) | 13(15) | 16(34) | 58(34) | 21(34) | 44(34) | 68(34) | 7(62) | 16(70) | 33(34) | 12(135) | 35(29) |
| Nanyuki county referral | 37(325) | 19(325) | 18(255) | 20(326) | 23(326) | 24(325) | 17(321) | 42(143) | 10(325) | 11(325) | 52(325) | 26(325) | 59(325) | 69(331) | 3(35) | 9(24) | 26(30) | 9(53) | 39(26) |
| Ruiru family clinic | 25(8) | 13(8) | 0 | 13(8) | 13(8) | 13(8) | 13(8) | 50(2) | 0 | 13(8) | 63(8) | 13(8) | 38(8) | 38(8) | 0 | 13(66) | 0 | 13(66) | 13(79) |
| MMM | 48(159) | 21(159) | 0 | 21(160) | 21(159) | 23(159) | 24(160) | 41(94) | 4(159) | 13(159) | 59(160) | 14(160) | 71(161) | 57(160) | 0 | 10(22) | 15(24) | 7(78) | 38(27) |

Key; AMC- Amoxicillin Clavulanic acid, AMP- Ampicillin, CXM-Cefixime, CAZ-Ceftazidime, CRO-Ceftriaxone, CTX-Cefotaxime, FOX-Cefoxitin, ATM-Aztreonam, AMK-Amkacin, GEN-Gentamicin, NAL-Nalidixic Acid, CIP-Ciprofloxacin, SMX-Sulfamethoxazole, TMP-Trimethoprim, FOS-Fosfomycin, NIT-Nitrofurantoin, LNZ-Linezolid, CHL-Chloramphenicol, TCY-Tetracycline, MMM-Mary Mother Mission Mukuru hospital, CMR-Centre for Microbiology Research.

**Fig 2:**
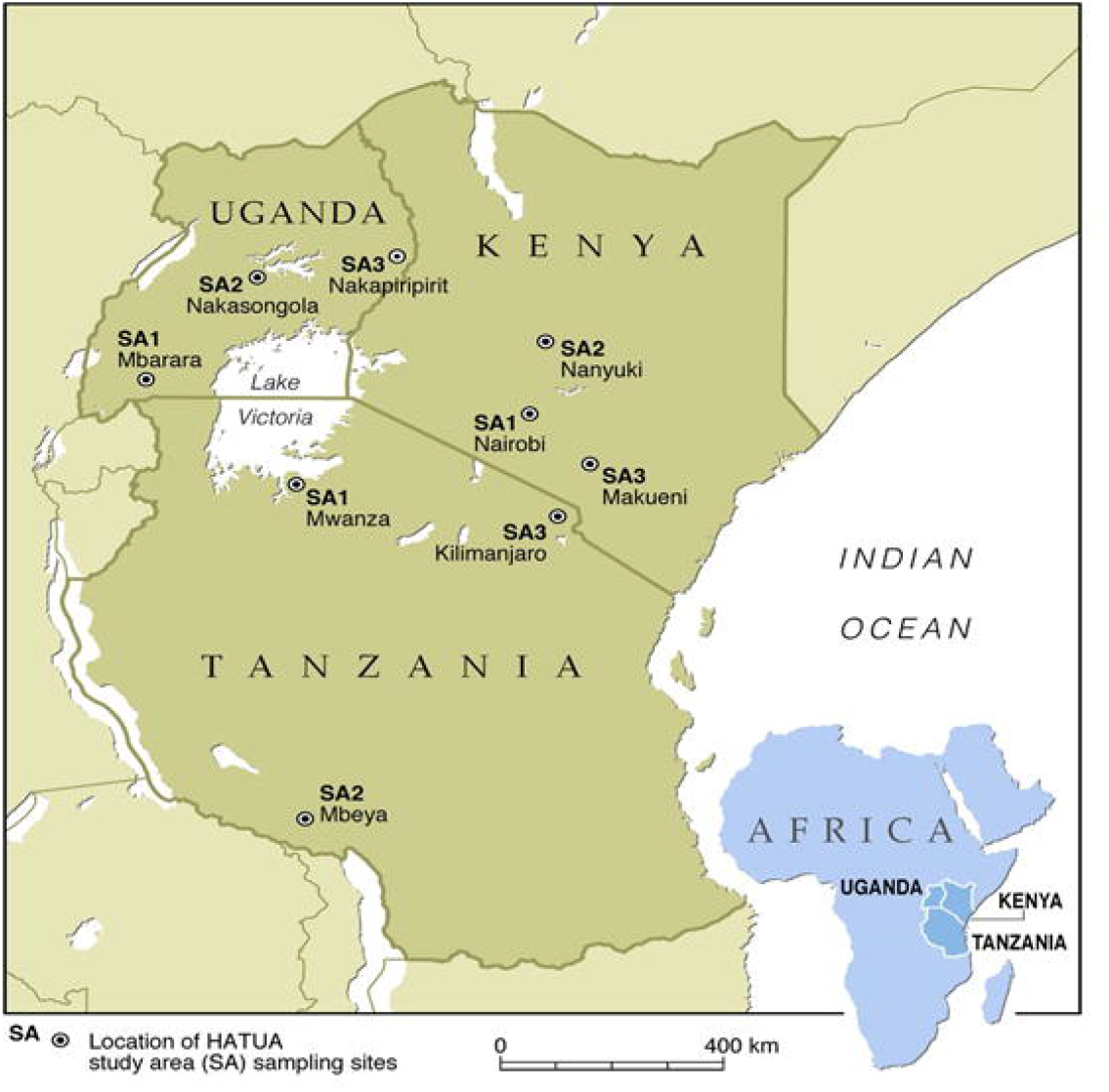
Percentage proportion of Uropathogens recovered from 1027 positive cultures. **Key; miscellaneous(*Serratia, Bacillus* and *Streptococcu*s ssp), E.coli-*Escherichia coli***

**Fig 3.**
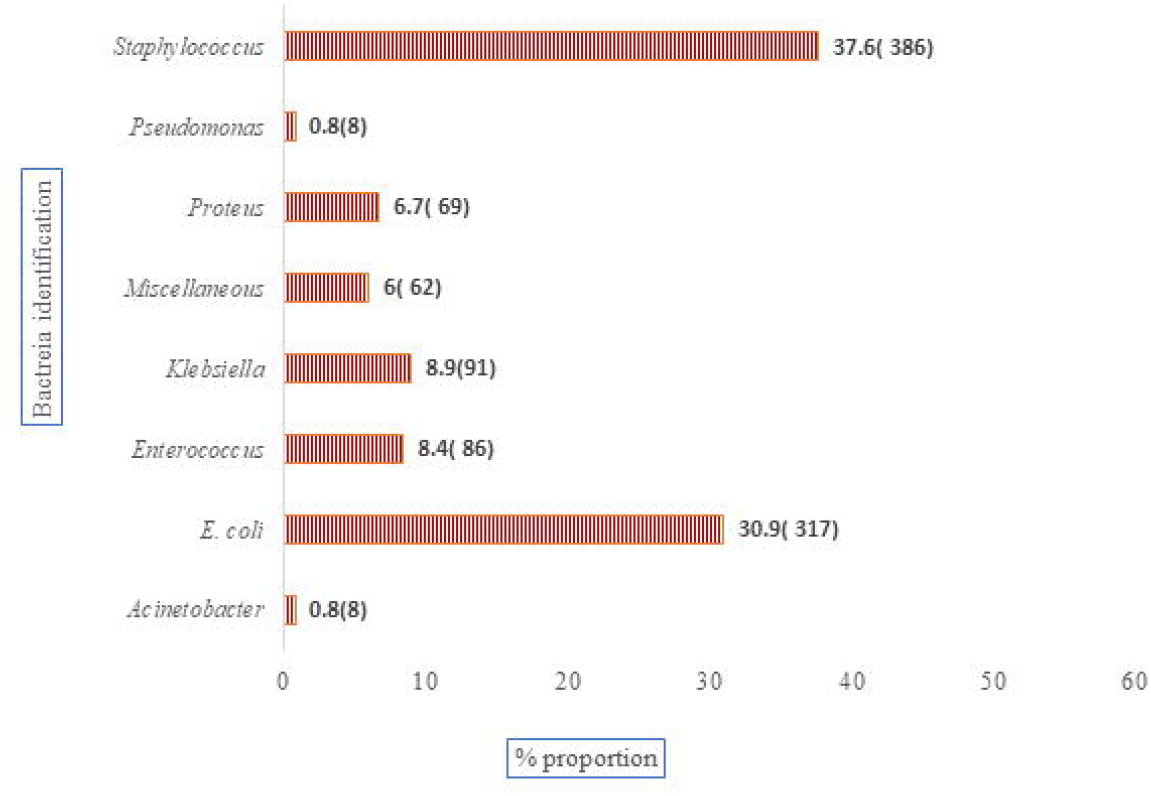
antibiotics resistance profiles (%) of all the uropathogens recovered: AMC- AMC- Amoxicillin Clavulanic acid, AMP- Ampicillin, CXM-Cefixime, CAZ-Ceftazidime, CRO-Ceftriaxone, CTX- Cefotaxime, FOX-Cefoxitin, ATM-Aztreonam, AMK-Amkacin, GEN-Gentamicin,NAL-Nalidixic Acid, CIP-Ciprofloxacin, SMX-Sulfamethoxazole, TMP-Trimethoprim, FOS-Fosfomycin, NIT-Nitrofurantoin, LNZ- Linezolid, CHL-Chloramphenicol, TCY-Tetracycline, Numbers in brackets are the n.

## Discussion

In the current study, the prevalence of UTI was 54.1%, where of the those with UTI, Gram-positive bacteria were more prevalent at 51% than Gram-negative (48%) and yeast at (2%). This prevalence is higher than that from a study by Donkor *et al*. (2019) in Accra, Ghana, who reported a prevalence of 10.1% from patients who were clinically suspected of having a UTI [17]. In neighboring Uganda and Tanzania, a prevalence of 32.2% and 28%, respectively, have been reported, which considerably varies from the 54.1% recorded in the current study [18][9]. This implies that the community-acquired UTI burden varies across geographical locations and regions and is affected by varying study designs.

Furthermore, Onyango *et al*. reported a prevalence of 15.7% in a cross-sectional study conducted among pregnant women in a maternity hospital in Kenya [8]. In contrast, UTI prevalence in the above African studies significantly varies from the 11% overall prevalence in the United States where the target population was older women, both pregnant and not pregnant [19]. The variation in UTIs across regions may be attributed to variations in sanitation standards, urine collection procedure and sample, UTI history, patients’ age, and sex composition across the populations. Moreover, the variation in prevalence may result from differences in patient and test inclusion criteria. For instance, the present study recruited patients who only presented with UTI-like symptoms. Therefore, their urine samples were cultured regardless of their nitrates and leukocyte esterases positivity rate. On the other hand, in the study by Odoki and colleagues, the recruited patients who presented with UTI-like symptoms had to cleanse the urethral area with castile soap towelette, reducing skin contamination chances [18]. This implies that there is need for more standardized testing for UTIs for better comparison and surveillance in future.

In the present study, Gram-positive uropathogens were more prevalent (51%) than Gram-negative uropathogens (48%). This contrasts with previous studies documenting a lower prevalence of Gram-positive bacterial isolates at 21% and 12%, respectively[8] [20]. The predominance of Gram-positive bacteria could imply changing patterns of uropathogens. Other possibilities could be contamination from the skin during urine collection.

Furthermore, the recovery of *Staphylococcus* spp. as the most dominant pathogen (34%) was different in other studies with *E. coli* as the main uropathogen. For example, in a study conducted in Kenya [20] the prevalence of *Staphylococcus* species was 21% and that of *E. coli* 38.5%, while another study conducted at Pumwani Hospital had a prevalence of 15.1% and *E. coli* at 44.5% [8]. Also, a study conducted in Pakistan resulted in a coagulase*-*negative staphylococci prevalence of 30.6% [21]. Additionally, *E. coli* was the second predominant uropathogen (26%), and the most prevalent Gram-negative bacteria, followed by *Klebsiella* spp. (9%) and *Proteus* spp. (7%) which is in agreement with a study carried out in Turkey that had *E. coli* as the most frequently isolated urinary pathogen (63.7%), followed by *K. pneumoniae* (18.7%) [22]. The recovery of *Pseudomonas* (0.7%) and *Acinetobacter* (1.3%), besides *E. coli* and *Klebsiella spp*., represents the most resistant bacterial populations to commonly used antibiotics, including carbapenems, and this poses a public health threat as there will be limited options to treat UTIs which are easily managed by antibiotics.

Antimicrobial resistance in these uropathogens may have had a significant implication in managing UTIs. For instance, the study shows a high prevalence of uropathogens with high resistance toward commonly UTI antibiotics, such as trimethoprim (64%), sulfamethoxazole (57%), and nalidixic acid (57%), which corroborates previous studies[8] [23]. Godman *et al. (*2018), shows that these are among the commonly prescribed antibiotics at most outpatient health facilities and are readily available over the counter at chemists and pharmacies[24]. The study found nitrofurantoin, amoxicillin-clavulanic acid, with the trade name Augmentin(tm), and cefepime as the better choice across all the study sites; this could be attributed to the fact that these drugs are not the first choice of treatment. The Gram-negative bacteria exhibited more resistance to the commonly used UTI antibiotics than the Gram-positive bacteria, which was in agreement with a recently published study in Kenya that had the same resistant trend in uropathogens isolated from pregnant women[8]. Resistance to ciprofloxacin ranged from 12 to 67%. This trend must be closely watched since fluoroquinolones are superior to trimethoprim/sulfamethoxazole (TMP/SMX; co-trimoxazole) for empirical therapy due to the relatively high prevalence of TMP/SMX resistance among uropathogens causing pyelonephritis [25][26]. Additionally, *Pseudomonas* spp. and *Acinetobacter* spp. revealed resistance towards nitrofurantoin at 38% and 25%, respectively. This is worrying as nitrofurantoin appears to be an ideal alternative to co-trimoxazole and fluoroquinolones for empirical treatment of uncomplicated UTIs, especially given the current prevalence of antibiotic resistance among community uropathogens [27].

Additionally, variation in resistance across recruitment sites was significant. This could be attributed to self-medication practices where the community members buy the currently available drugs from the chemists around[28]. Self-medicating individuals also predominantly use the same affordable antibiotics[28]. However, different areas have a diverse supply of antibiotics hence the variation in resistance[29]. This suggests limited treatment options, making UTIs challenging to treat and posing an apparent threat to public health. Elsewhere, the high rate of resistance of uropathogens to commonly used antibiotics and the emergence of *S. aureus* as a significant causative agent could indicate a problem at the community or facility level due to wrong prescription, over-prescription, or poor adherence. This highlights the need for continuous local surveillance of susceptibility patterns of uropathogens so that a guide for empirical antibiotic prescription can be updated.

## Conclusion

The study documents a high prevalence of UTIs and Gram-positive bacteria as the most dominant uropathogens implying changes in uropathogens patterns. The possible consequence of this could be a wrong antibiotics prescription since Gram-negative bacteria have previously been prevalent. Furthermore, high resistance rates towards quinolones, sulfamethoxazole, and trimethoprim were reported, which are the commonly used drugs as they are cheap to buy and readily available. These findings, therefore, indicate the need to for more robust standardised surveillance is needed to confirm the patterns observed. The study also highlights the need for the identification of the causative agent of UTI, even though not all hospitals have the capacity to carry out the cultures. Therefore, annual surveillance of the circulating UTI causative agents should be adopted to update the national guidelines for antibiotics empirical therapy accordingly.

## Data Availability

All data produced in the present study are available upon reasonable request to the authors

## Study limitations

This study recruited patients seeking treatment for UTI-like symptoms and therefore prevalence reported in the present study may not reflect the actual prevalence in the community. This assumption is further reinforced by the fact a substantial number of patients in poor resource settings frequently opt for self-treatment and only visit hospital for severe UTI cases or when symptoms have persisted[30]. Regardless, our study provides a good glimpse into the UTI burden in Kenya, considering this is the largest UTI study in Kenya by sampling sites and sample size.

## Authors contribution

Susan Kiiru led the antibiotic susceptibility testing team and has drafted this manuscript. In addition, John Maina and Japhet Katana were involved in the crucial components of the study’s methodology formulation and actualization. Dr. John Mwaniki was involved in the conceptualization and actualization. of the study, and reviewed and edited the initial drafts of this paper. Katherine Keenan, Stephen H. Gillespie, and John Stelling contributed to the study design and protocol writing and reviewed the manuscript. Dr. John Kiiru is part of the team that conceptualized the study, and the HATUA Kenyan Chapter lead and provided overall supervision of the study. Prof. Matthew Holden is the HATUA principal investigator and the one who received the funding.

## Acknowledgments

We are indebted to the Medical Research Council UK for providing financial support to the study. We also acknowledge the HATUA study collaborators, St. Andrews University, Scotland, Makerere University, Uganda, and the Catholic University of Health and Allied Sciences in Mwanza, Tanzania. We also appreciate the health facilities that allowed us to conduct our study on their premises. Finally, our heart goes to the participants, whom the study couldn’t have been possible without them.

